# Implementation of risk prediction models using electronic health data for early detection of pancreatic cancer: a prospective pilot feasibility study

**DOI:** 10.64898/2026.09.22.26363704

**Authors:** Bechien U. Wu, Tiffany Q. Luong, Eva Lustigova, Christie Jeon, Eric J. Puttock, Rebecca H. Moon, Mercedes A. Munis, Wansu Chen

**Affiliations:** Department of Research & Evaluation, Southern California Permanente Medical Group, Pasadena, CA; Department of Gastroenterology, Kaiser Permanente Los Angeles Medical Center, Los Angeles, CA; Tulane Center for Clinical Research, Tulane University School of Medicine, New Orleans, LA; Department of Epidemiology, UCLA Fielding School of Public Health, Los Angeles, CA

## Abstract

**Background & Aims:** We previously developed multiple machine-learning and regression-based risk prediction models using retrospective electronic health records (EHR) to estimate near-term risk of pancreatic cancer (PC). This pilot study aimed to assess the feasibility of an early detection strategy combining initial EHR-based risk prediction with downstream imaging and blood-based testing.

**Methods:** This prospective study included adults aged 50–84 years with an estimated ≥1% 18-month risk of PC, identified between January 2021 and March 2023 using four previously validated models. Patients with a history or active suspicion of PC were excluded. Participants completed cross-sectional imaging and CA19-9 labs at baseline and again at 18 months. Outcomes included enrollment rates, test completion rate, and burden of incidental findings.

**Results:** Of 1,251 eligible patients, 102 (8.2%) enrolled (median age 69 years [IQR 62-75]; 58% women; 48% Hispanic, 34% White, 11% Black, and 7% Asian). At baseline, 89 (87%) completed imaging and 98 (96%) completed CA19-9. At 18 months, 79% completed both imaging and CA19-9. Ten (10%) participants underwent additional clinical evaluations triggered by imaging findings. Evaluations included an endoscopic ultrasound, renal imaging, and FibroScan. CA19-9 results were normal in most participants, though 18% had elevated levels at baseline.

**Conclusions:** Real-time EHR-based ML algorithms can be combined with blood- and imaging-based surveillance for early detection of PC with acceptable burden of incidental findings. However, improved patient engagement is a key aspect that will need to be addressed in order to successfully scale an AI-guided strategy for early detection.

## Introduction

Despite a relatively low incidence of approximately 14 cases per 100,000 persons,^1^ pancreatic cancer (PC) is the third leading cause of cancer-related death in the United States.^2^ The poor prognosis of PC is primarily due to being diagnosed at an advanced stage, often with distant metastases when treatment options are limited. Early detection has the potential to improve survival; however, the United Services Preventative Services Task Force does not recommend population-based screening due to limited effectiveness.^3^ This has created an unmet need for targeted screening strategies capable of identifying high-risk individuals before symptoms develop.

Recent advancements in electronic health records (EHR) and artificial intelligence (AI) have created opportunities for innovative early detection strategies. Machine learning (ML) algorithms trained on large-scale EHR data have shown promising accuracy in identifying individuals at elevated risk for PC, in both general and high-risk populations.^4–11^ These algorithms leverage routinely collected clinical variables, such as age, laboratory values, weight change, and symptoms, to estimate near-term cancer risk.

To efficiently identify patients with early-stage pancreatic cancer, we and others have proposed a targeted approach that involves a systematic search for early warning signs in the electronic health record followed by a focused clinical evaluation.^12^ This framework, which has been referred to as “heuriskance” lends itself to application of risk prediction models as the initial step for risk-stratification to identify patients at near term risk of developing PC followed by subsequent focused clinical evaluation.

In this prospective pilot study, we aimed to assess the real-world feasibility of integrating EHR-based ML models with downstream diagnostic testing, including blood-based biomarkers and cross-sectional imaging, for the early detection of PC. Specifically, we applied four previously validated prediction models to identify patients at elevated risk for pancreatic cancer within an integrated healthcare system and invited those individuals to participate in a structured diagnostic protocol. By evaluating patient identification, recruitment, testing adherence, diagnostic yield, and the management of incidental findings, we sought to gain insights into both the clinical and operational dimensions of implementing AI-driven screening strategies.

To our knowledge, this study represents one of the first prospective evaluations of ML-based PC risk models embedded within clinical care pathways. By combining algorithmic risk stratification with subsequent diagnostic testing, we aim to inform the design of future large-scale screening initiatives and help accelerate the safe, effective translation of AI tools into real-world oncology practice.

## Methods

Study Design and Setting: We conducted a pilot prospective cohort study from January 2021 to January 2025 in Kaiser Permanente Southern California (KPSC), an integrated health system that provides comprehensive care for approximately 4.9 million members in Southern California. The study was approved by KPSC’s Institutional Review Board (Protocol #12135) and was registered at ClinicalTrials.gov (NCT04883450). The study is reported in accordance with the DECIDE-AI framework.^13^

Participant Eligibility: Eligible patients were adults aged 50 to 84 years who did not have a prior history or active suspicion of PC, any other active malignancy, end-stage renal disease, or were not pregnant. Patients who had been residents of a skilled nursing facility or hospice within the past year were also excluded, as were those unable to provide informed consent in either English or Spanish. To ensure the necessary data for risk scoring, eligible patients were required to have at least one of the following measurements recorded within six months prior to their eligibility date: weight, hemoglobin A1c (HbA1c), and alanine aminotransferase (ALT).

Previously Validated Models: The study utilized four previously developed PC risk prediction models (**Supplemental Table 1)**. These included two random survival forest (RSF) models, an extreme gradient boosting [XGB] model, and a Cox proportional hazards regression [COX] model. The second RSF model, referred to as the early detection RSF (eRSF) model, was developed from a study population with ≥90 days of cancer-free follow-up, in an effort to identify parameters associated with earlier stage of pancreatic cancer.^8^ All four models were developed using data from KPSC and were externally validated on data from the Veterans Affairs health systems.^8,14^ These models use routinely collected clinical parameters including age, weight, HbA1c, ALT and abdominal pain. All model parameters were extracted electronically from the EHR as previously described.^8^

Model Implementation: The validated models were implemented in three phases. During Phase 1 (January 29 to October 19, 2021), we implemented the first RSF model. In Phase 2 (November 16, 2021, to December 13, 2022), we applied the eRSF model. Phase 3 (January 10 to March 5, 2023) involved the simultaneous application of all four models. The rationale for this approach was to first determine the feasibility of prospective implementation using a single modeling approach. Subsequently, we evaluated gains and challenges associated with simultaneous application of additional models. Models were applied in real-time at two-week intervals to identify patients with a predicted PC risk ≥1% within 18 months (**Supplemental Figure 1**).

Recruitment and Informed Consent: Potential eligible patients based on model implementation were further screened by trained research associates to confirm eligibility for study recruitment. Additionally, for each eligible patient, their primary care physician was notified of the study and the patient’s eligibility. Physicians were provided an opportunity to opt-out on behalf of their patients, inquire further about the study, or express any study-related concerns. Patients were contacted directly by the study team through a combination of emails, letters, and phone calls for study participation if no physician opt-out was received after 3 days. Informed consent was obtained from all participants prior to the initiation of any study-related procedures. As a demonstration of feasibility of the early-stage clinical screening, targeted accrual (sample size) was set at 100 participants.

Study Procedures: Participants who provided informed consent were invited to undergo a series of assessments and tests for early detection of PC. At baseline, participants were invited to complete a carbohydrate antigen 19-9 (CA 19-9) test and a magnetic resonance imaging (MRI) of the pancreas. Testing for CA 19-9 used the Roche Elecsys® assay, and the MRI used 3-Tesla scanners with a standard pancreas protocol. For participants unable to complete an MRI (e.g., due to claustrophobia), a contrast-enhanced computed tomography with multi-detector pancreas protocol (MDCT) was offered as an alternative. Participants were also asked to complete a follow-up CA 19-9 test and imaging at 18 months.

Outcomes Measures: The primary outcome for this study was feasibility, measured as the proportion of participants who successfully completed at least one imaging study (MRI or MDCT) or one CA 19-9 test. Secondary outcomes included (1) yield of testing measured as positive predictive value (PPV) for PC, and (2) the acceptability of the screening strategy, assessed by the accrual rate, defined as the proportion of eligible patients who enrolled. PC diagnosis was assessed through review of participants’ EHR. The frequency and nature of additional clinical evaluations resulting from imaging findings obtained as a result of study participation were also monitored. Extra-pancreatic findings were categorized according to standards previously established for CT colonography^15^ (see **Supplemental Table 2**) with only E3-E4 findings reported.

Safety was assessed by tracking any adverse events related to study procedures, including difficulties in completing imaging studies. Provider concerns were assessed on the frequency of physician opt-out. Finally, to identify potential bias in study participation, the demographic characteristics of study participants (age, sex, and race/ethnicity) were compared to those of the overall eligible population identified as having a ≥1% predicted risk of PC over 18 months.

Statistical Analysis: Descriptive statistics were used to summarize the characteristics of the study population and study outcomes. Continuous variables were reported as means with standard deviations or medians with interquartile ranges, while categorical variables were presented as frequencies and percentages.

The study was not designed or powered to assess cancer detection efficacy.

## Results

A total of 1,653 patients with a predicted risk of ≥1% for developing PC and meeting inclusion/exclusion criteria were identified during the study period. Among these, 1,288 (77.9%) were confirmed as eligible for recruitment after manual review of patient charts, with reasons for exclusion reported in **Supplemental Table 3**. Among the confirmed eligible patients, 20 (1.6%) were excluded at the request of their primary care physicians. The cited reasons for physician opt-out are presented in **Supplemental Figure 2**. An additional 17 (1.3%) patients were not contacted for study participation as the recruitment target was achieved prior to outreach.

A total of 102 patients (8.2% of the 1,251 patients contacted) consented and were enrolled in the study. A flow diagram depicting cohort assembly is presented in **Figure 1**. Among the 805 patients who declined participation, the most common reason cited was lack of interest (39.0%), followed by excessive distance to study site (16.3%), poor health (11.8%) and lack of time (11.1%) (**Figure 2**).

**Figure 1.**
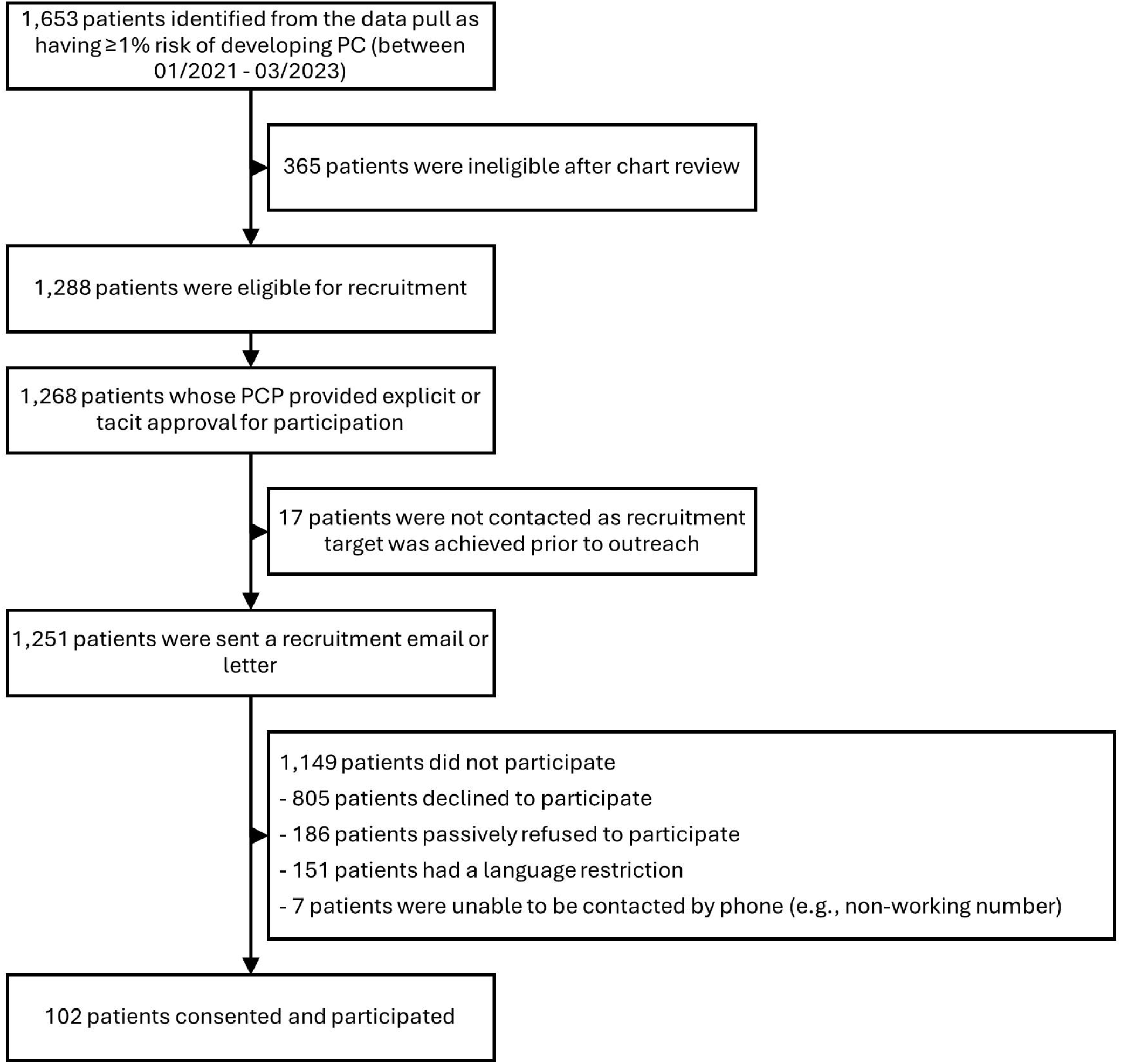
Flow diagram of study recruitment.

**Figure 2.**
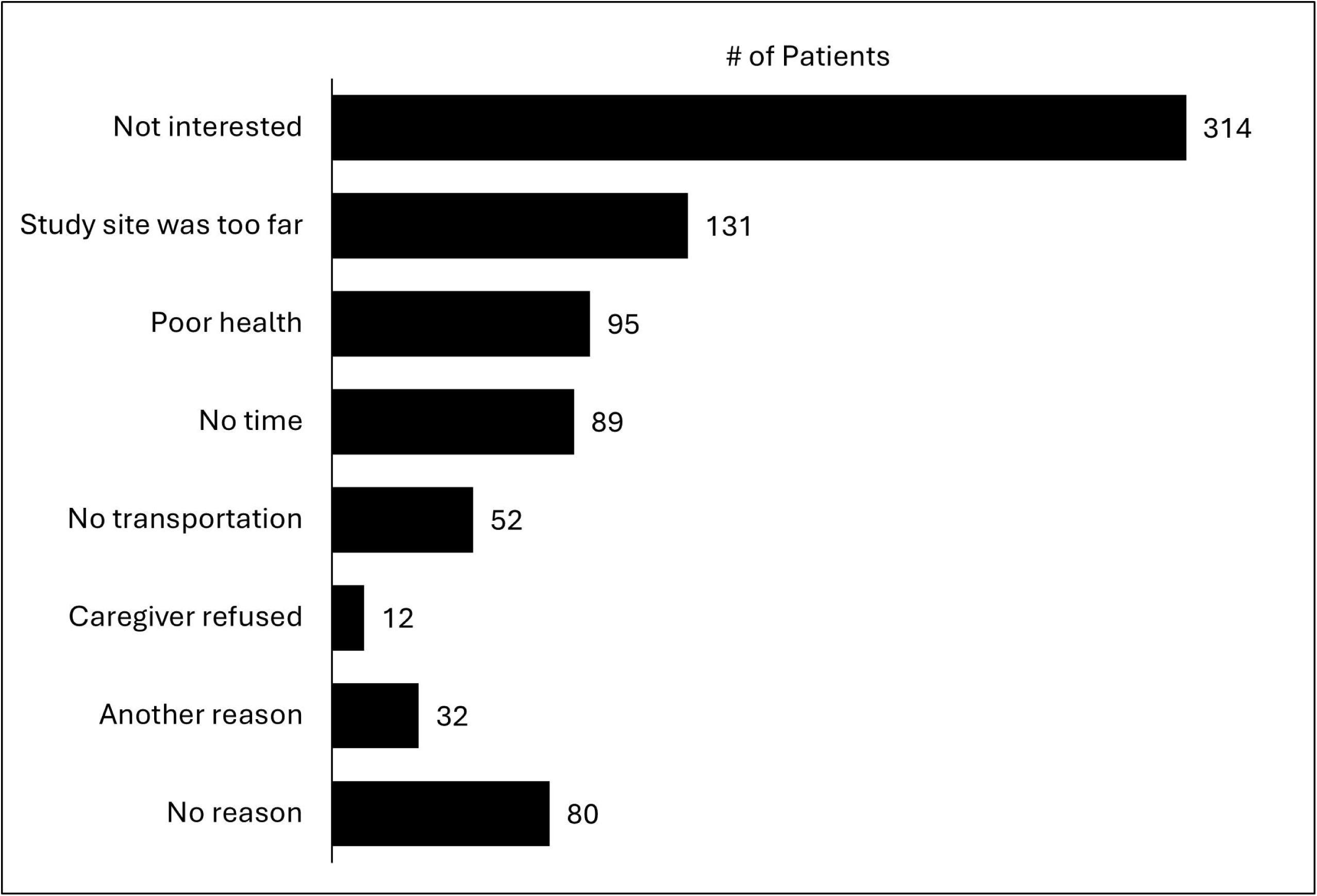
Primary reasons for declining to participate, among 805 patients who declined.

The median age of study participants was 68.8 years (IQR 61.6-75.3), and 57.8% were female. In terms of race and ethnicity, 48.0% participants identified as Hispanic, 34.3% as non-Hispanic White, 10.8% as non-Hispanic Black and 6.9% as non-Hispanic Asian or Pacific Islander. Compared to the eligible non-participating population, study participants were on average younger (mean age 68.7 vs. 72.6 years, p<0.0001), included a higher proportion of White individuals (34.3% vs. 22.4%, p<0.01), and were more likely to speak English (86.3% vs. 69.8%, p<0.001). The distribution by patient sex was similar between participants and non-participants (57.8% vs. 52.2% female, p=0.27). Detailed demographic characteristics are summarized in **Table 1**.

**Table 1.**
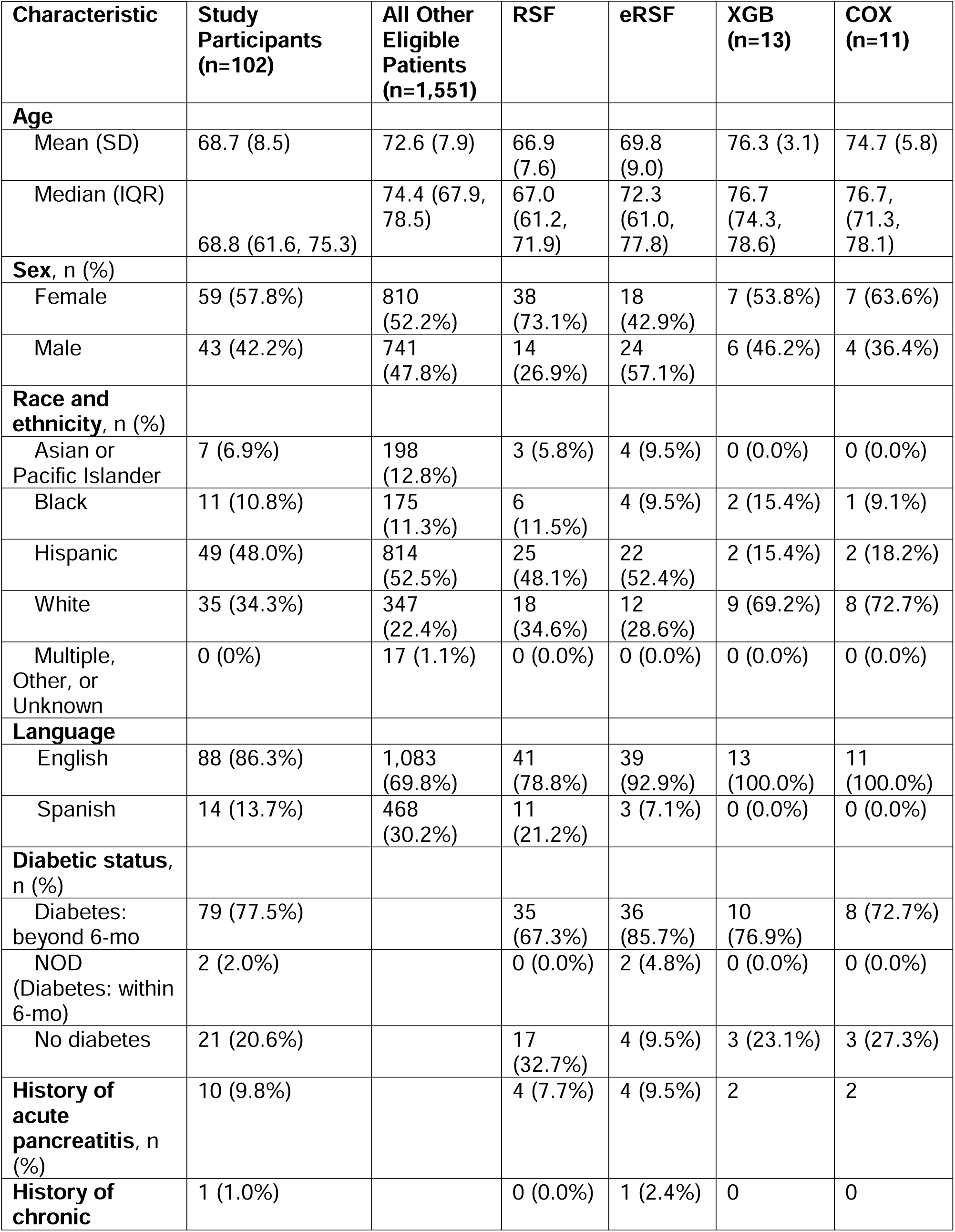
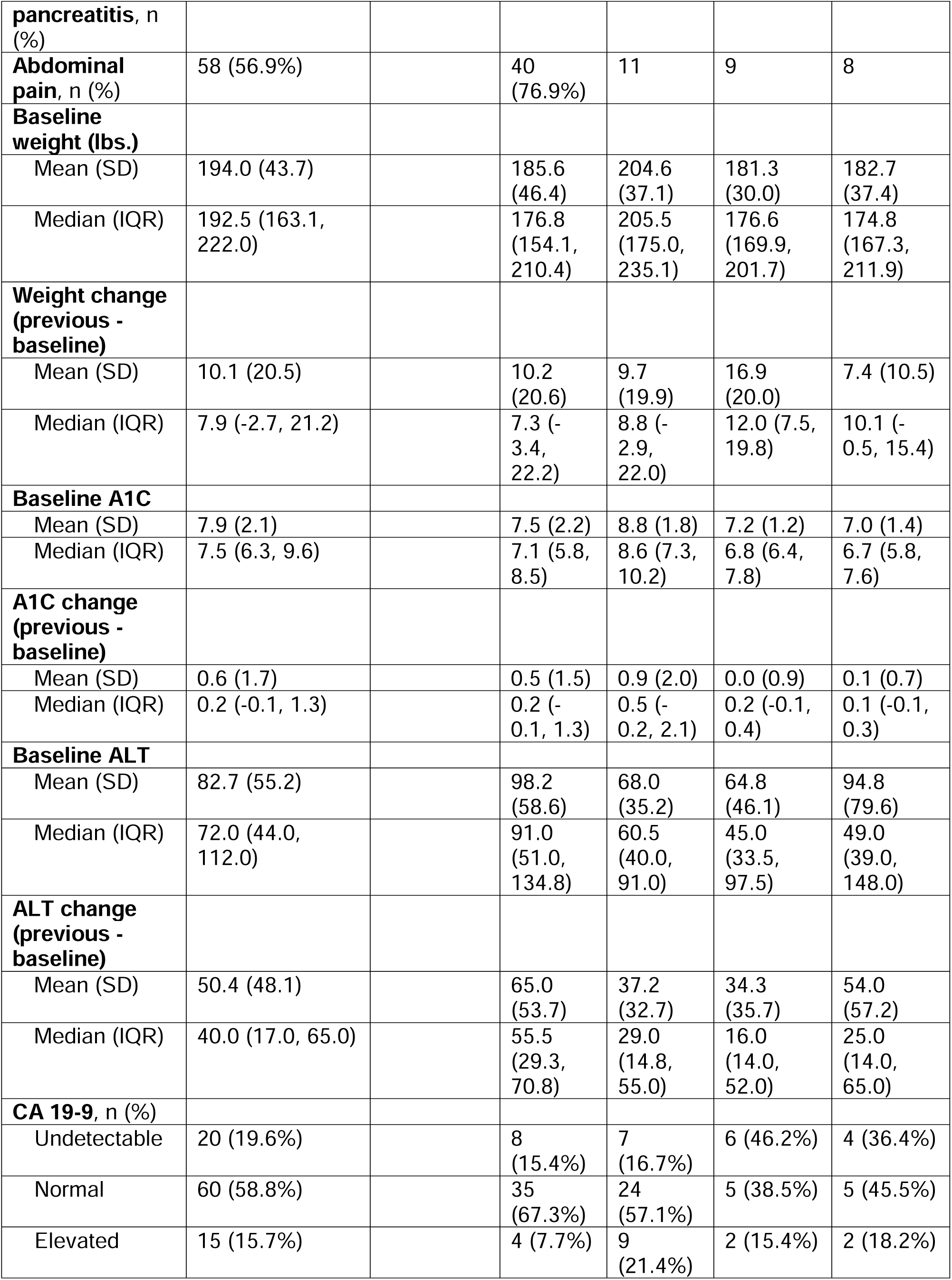
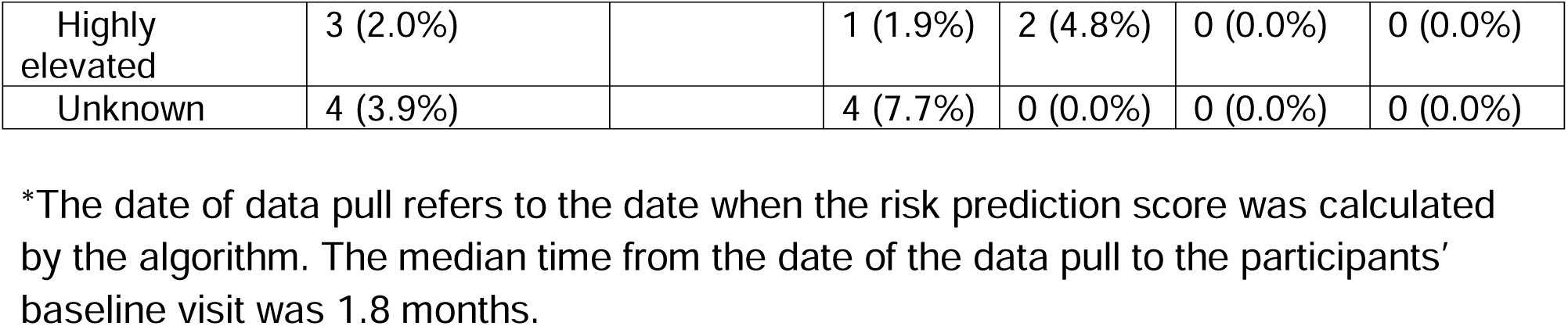
Participant and cohort characteristics at the time of the data pull* (n=1,653).

### Model Implementation

The previously described models were prospectively applied to identify patients at elevated risk for PC. During the entire study period, the models identified a total of 1,653 high-risk patients, of whom 1,251 were contacted and 102 (8.2%) ultimately enrolled (**Table 2**). During Phase 3 (January 10 – March 5, 2023), when all four models were applied simultaneously, 362 patients were identified as high-risk, and 270 were contacted (**Table 2**). Among these, 14 participants (5.2%) were enrolled. The Venn diagrams presented in **Figure 3** pertain specifically to this final phase and illustrate the overlap and distinctiveness of patient identification across models. While some patients were flagged by multiple models, each model also identified unique patients. Most notably, XGB and Cox each contributed substantial numbers of high-risk patients who were not identified by the RSF models. Among the 14 participants enrolled in Phase 3, the majority (13) were identified by XGB, underscoring potential differences in the risk profiles prioritized by each algorithm.

**Figure 3.**
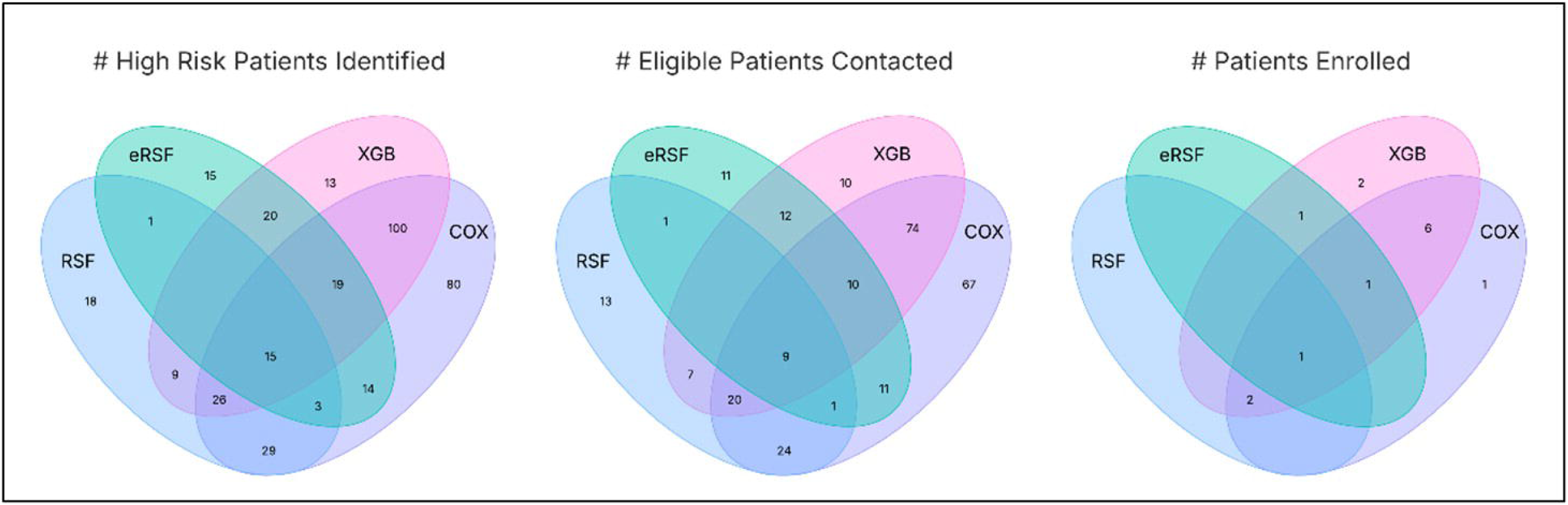
Venn diagrams illustrating the number of high-patients identified, contacted, and enrolled, by each risk prediction model, during Phase 3 (January 10 – March 5, 2023) of the study. Footnote for Figure 3: COX, Cox proportional hazards regression; eRSF, early detection random survival forest; RSF, random survival forest; XGB, extreme gradient boosting.

**Table 2.** Number of high-risk, eligible, and enrolled patients by model.

|  | Models Applied* |  |  |  |  |
| --- | --- | --- | --- | --- | --- |
|  | RSF | eRSF | XGB | Cox | Total |
| **Entire Study Period:**<br>**January 29, 2021 – March 5, 2023** |  |  |  |  |  |
| # High Risk Patients Identified | 707 | 772 | 202 | 286 | 1653 |
| # Eligible Patients Contacted | 551 | 560 | 142 | 216 | 1251 |
| # Patients Enrolled (%)** | 52 (9.4) | 42 (7.5) | 13 (9.2) | 11 (5.1) | 102 (8.2) |
| **Phase 3:**<br>**January 10 – March 5, 2023** |  |  |  |  |  |
| # High Risk Patients Identified | 101 | 87 | 202 | 286 | 362 |
| # Eligible Patients Contacted | 75 | 55 | 142 | 216 | 270 |
| # Patients Enrolled (%) | 3 (4.0) | 3 (5.5) | 13 (9.2) | 11 (5.1) | 14 (5.2) |

COX, Cox proportional hazards regression; eRSF, early detection random survival forest; RSF, random survival forest; XGB, extreme gradient boosting.
\*Numbers are not mutually exclusive between models, as a patient may be identified as high risk by one or multiple models.
\*\*The proportion of enrolled patients is calculated among the eligible patients contacted.

### Completion Rates

Among the 102 enrolled participants, imaging completion rate was 87% (including either MRI or CT) with 96% completion of the CA 19-9 blood test at baseline. At the 18-month follow-up, 79% of participants completed imaging and 79% completed the CA 19-9 test (**Table 3**). The median time from baseline to completion of imaging was 1.8 months, while the median time to complete CA 19-9 testing was 1.5 months. Detailed completion rates at each follow-up interval are presented in **Table 3**.

**Table 3.** Completion rates of the imaging and CA 19-9 lab, and time to completion of the imaging and CA 19-9 labs (n=102).

| Study Task | Value |
| --- | --- |
| **Baseline** |  |
| Imaging | 89 (87%) |
| *Computed Tomography* | 3 (*includes 2 research, 1 clinical*) |
| *Magnetic Resonance Imaging* | 86 (*includes 85 research, 1 clinical*) |
| Median (IQR) Months to Imaging* | 1.8 (1.3, 2.5) |
| CA 19-9 Lab | 98 (96%) |
| CA 19-9 Lab Result |  |
| *Undetectable* | 20 |
| *Normal ($\leq 35$ U/mL)* | 60 |
| *Elevated ($>35$ - $<100$ U/mL)* | 15 |
| *Highly Elevated ($\geq 100$ U/mL)* | 3 |
| *Missing* | 4 |
| Median (IQR) Months to CA 19-9 Lab* | 1.5 (0.8, 2.6) |
| **18-Month** |  |
| Imaging | 81 (79%) |
| *Computed Tomography* | 9 |
| *Magnetic Resonance Imaging* | 72 |
| Median (IQR) Months to Imaging* | 17.7 (16.3, 19.3) |
| CA 19-9 Lab | 81 (79%) |
| CA 19-9 Lab Result |  |
| *Undetectable* | 32 |
| *Normal ($\leq 35$ U/mL)* | 34 |
| *Elevated ($>35$ - $<100$ U/mL)* | 13 |
| *Highly Elevated ($\geq 100$ U/mL)* | 2 |
| *Missing* | 21 |
| Median (IQR) Months to CA 19-9 Lab* | 17.2 (16.5, 18.2) |

\*Time is calculated from the participants' baseline visit.

### Imaging Findings and Clinical Evaluations

Among the 102 study participants, 89 completed the baseline imaging and 81 completed the 18-month imaging (**Table 3**). Imaging performed at baseline identified various pancreas findings, including pancreatic cysts (n=3), pancreatic duct dilation (n=2), and one lesion that was subsequently determined to be PC (**Table 4**). At 18 months, imaging revealed additional pancreatic cysts (n=19) in addition to the PC case that was diagnosed. The PC case was noted to have a 2 cm pancreas lesion on baseline imaging. Initial endoscopic ultrasound testing with fine needle aspiration at that time was non-diagnostic. A diagnosis of PC (borderline resectable) was made prior to the scheduled 18-month follow-up based on repeat clinical testing. A summary of the pancreatic and extra-pancreatic findings is presented in **Table 4**. Overall, 9 (10%) patients underwent additional clinical testing as a result of the baseline MRI (6 pancreas-related testing, 3 extra-pancreatic). A total of one pancreatic cancer case was detected in the study population, resulting in a positive predictive value of 0.98%.

**Table 4.** Summary of imaging findings and clinical evaluations performed among study participants.

| **Timepoint** | **Location** | **Imaging Findings** | **Clinical Evaluations\*** |
| --- | --- | --- | --- |
| **Baseline** | Pancreas | • Pancreatic cysts (n=3)**<br>• Pancreatic duct dilatation (n=2)<br>• Chronic pancreatitis (n=1)<br>• Pancreatic cancer (n=1) | • Endoscopic ultrasound (n=6) |
|  | Extra Pancreatic | • Kidney mass (n=1)<br>• Renal cyst (n=1)<br>• Fatty liver (n=34) | • Esophagogastroduodenoscopy (n=1)<br>• FibroScan (n=1)<br>• Renal ultrasound (n=1)<br>• Renal biopsy (n=1) |
| **18-Month** | Pancreas | • Pancreatic cysts (n=19)***<br>• Pancreatic duct dilatation (n=3) | • CT (n=2)<br>• Endoscopic ultrasound (n=1) |
|  | Extra Pancreatic | • Renal cyst (n=18)<br>• Hepatic cyst (n=11)<br>• Fatty liver (n=29) | • CT (n=6)<br>• MRI (n=3) |

\*The numbers in parentheses refer to the number of patients with the finding or who underwent the clinical evaluation. Numbers are not mutually exclusive.
\*\*Sizes of the cysts ranged between 12 – 20 mm.
\*\*\*Sizes of the cysts ranged between 3 – 18 mm

### CA 19-9 Performance

At baseline, 98 participants (96%) completed CA 19-9 testing. Among these, 20 (20.4%) participants had undetectable CA 19-9 levels, 60 (61.2%) were within the normal range (≤35 U/mL), 16 (16.3%) were elevated (>35 to <100 U/mL), and two (2.0%) were highly elevated (≥100 U/mL) (**Table 3**). At 18 months, 81 participants (79%) completed CA 19-9 testing, with a similar distribution of results. Of note, the CA 19-9 for the participant later diagnosed with PC was in the normal range at baseline, and was elevated at the time of diagnosis.

### Safety and Adverse Events

There were no adverse events reported in relation to the imaging procedures or biospecimen collection. Participant tolerance for MRI was high, with minimal (n=3) participants opting for MDCT due to claustrophobia or contraindications for MRI.

## Discussion

In this prospective pilot study, we assessed the feasibility of a strategy that integrated ML and regression-based models with blood-based testing and cross-sectional imaging for early detection of PC in a real-world clinical setting. Our findings highlight both the promise and challenges of using AI algorithms for PC screening, offering valuable insights for future large-scale implementation.

We implemented four risk prediction models to identify patients at elevated risk for PC. These machine learning models were originally derived on data from KPSC from 2008-2017 using random survival forest (RSF), extreme gradient boosting as well as traditional cox proportional hazards regression with accuracy (AUC) ranging from 0.77 (95% CI 0.74-0.79), 0.78 (95% CI 0.76-0.80) and 0.74 (0.74-0.79) in the internal validation datasets, respectively.^14^ The model variables include age, abdominal pain, weight change, alanine transaminase (ALT) as well as glycated hemoglobin (A1c). We also applied a previously developed “early detection” model using RSF that only included data limited to 90 days prior to a diagnosis of PC.^8^ Further details regarding model development and validation can be found in the supplementary material. While numerous risk-prediction models have now been developed for early detection in PC,^4,5^ a coherent strategy that incorporates model generated risk for intervention has yet to be established.

The current study sought to determine the feasibility of blood-based testing as well as cross-sectional imaging based on model-predicted risk. Overall, a reasonably high completion rate of baseline testing (87% for imaging and 96% for CA 19-9) underscores the potential feasibility of this combined approach for early detection. However, the low overall accrual rate (8.2%) highlights potential challenges with future implementation with respect to patient engagement. Although the study was not powered to assess the performance of the prediction models, the observed PPV of 0.98% (was consistent with the *a priori* targeted threshold.

The current study advances previous work on prediction models for pancreatic cancer in several important ways. First, unlike most existing risk prediction models that have been developed and validated retrospectively, we applied our models prospectively in the context of real-time clinical practice. While previous evaluations have focused on measures of model performance, this real-time evaluation allowed us to shift attention to address complexities inherent in clinical implementation, including data availability, patient engagement, and the operational challenges of integrating risk prediction algorithms into further testing within the context of routine care. Second, our study is among the few to use multiple models concurrently, which allowed us to identify a broader range of high-risk patients. Different models may identify different high-risk individuals due to variations in their underlying algorithms and input variables, and this approach enabled us to capture an expanded high-risk cohort. Finally, the current study also demonstrates the feasibility of an integrated approach that combines blood-based testing (CA 19-9) as well as cross-sectional imaging (MRI as well as CT) to facilitate timely cancer detection among individuals identified as having increased risk by an EHR-based prediction model.

The performance of CA 19-9 in our cohort underscores both its limitations and the need for improved biomarkers for early detection of PC. The observed rate of undetectable CA 19-9 (19.6%)^16^ is consistent with previous clinical studies^16^ that have included Hispanic and non-Hispanic Black populations in the U.S.^17^ While it is unclear what proportion is attributable to the Lewis antigen null genotype, such a degree of uninformative results limits the utility of CA 19-9 as a standalone biomarker. However, several emerging blood-based tests have been shown to outperform CA 19-9 for early detection of PC including markers based on cell-free DNA,^18^ immune profiling^17^ as well as glycans of CA 19-9.^17^ The high completion rate of blood tests (96%) in our study is encouraging and suggests that future strategies integrating novel biomarkers could be well-accepted in clinical settings.

Our study revealed significant challenges related to patient engagement. Only 8.2% of eligible patients enrolled in the screening program, a rate substantially lower than those reported in established cancer screening efforts, such as lung, breast, or colorectal cancer screening, where participation typically ranges from 27% to 84%.^19,20^ Most notably, rates for lung cancer screening, which also makes use of cross-sectional imaging, have achieved rates of 25% among eligible adults.^21^ This comparatively low participation rate for patients with predicted near-term PC risk may be attributable, in part, to the research nature of our study. Notably, the most commonly cited reason for declining participation was lack of interest, followed by logistical barriers such as travel distance and time constraints. These findings raise important questions about how to optimize participation in AI-driven screening programs. Enhancing engagement of primary care providers, expanding access through convenient or at-home testing options, and using digital platforms to deliver education and reminders may improve uptake. Clear, accessible messaging that communicates the benefits of early detection and the minimal risks involved in screening will also be essential.

Our study also highlighted the challenge of managing incidental imaging findings, a well-recognized issue in screening programs that rely on cross-sectional imaging. In our cohort, 10% of participants underwent additional clinical evaluations based on imaging findings, including evaluations for pancreatic cysts, renal cysts, and fatty liver. This observed rate is consistent with other image-based screening programs that have applied a standardized classification system for incidental findings.^22^ Understanding the frequency of the incidental findings that potentially trigger additional workup is critical for evaluating the burden and cost-effectiveness of screening strategies. Future efforts to enhance model specificity, possibly through the integration of liquid biomarkers, could help further reduce the burden of unnecessary scans and incidental findings.

This study has several limitations. First, the relatively small sample size reflects the pilot nature of the study and limits the generalizability of the findings. Second, the low participation rate highlights the need for more effective patient engagement strategies, as previously discussed. Third, reliance on MRI-based imaging may limit the ability to scale this approach more broadly due to resource constraints. Fourth, the use of CA 19-9 as the primary blood-based marker for PC has known limitations in terms of sensitivity and specificity. The purpose of inclusion in the present study was to demonstrate the feasibility of incorporating blood-based testing as part of a more comprehensive early detection strategy. Future studies will likely benefit from incorporating more advanced blood-based biomarkers or multi-omics approaches as an intermediate step to identify high-risk patients to undergo subsequent cross-sectional imaging, potentially reducing unnecessary testing. Finally, the initial study start-up and recruitment efforts were hampered by the impact of post-COVID 19 re-opening of the healthcare system, which ultimately led to longer than anticipated period for phase 1 and 2, which limited the duration of phase 3 when all models were run simultaneously. We were therefore unable to draw meaningful conclusions regarding any differences with respect to model performance.

In conclusion, we have completed a prospective feasibility study that reflects many of the processes involved in early-stage live clinical evaluation of a ML approach for early detection in PC. Specifically, we piloted an approach that incorporates automated, real-time analysis of data from the electronic health record as well as subsequent blood-based testing and cross-sectional imaging. We identified several real-world challenges that include ensuring adequate support infrastructure for coordinating laboratory testing, managing imaging findings, and maintaining effective communication with patients and healthcare providers. Addressing these barriers will be a key step for successful large-scale implementation of ML algorithms for early detection of PC.

## Data Availability

Anonymized data that support the findings of this study can be made available from the corresponding author on reasonable request from qualified researchers with documented evidence of training for human subjects' protections.

## Acknowledgements

The authors thank the patients of Kaiser Permanente for helping to improve care through the use of information collected through our electronic health record systems. We also thank our research coordinators, Devon Luther and Kush Yadav, for their work in conducting the research visits, Rebecca Butler for her work in developing the risk prediction algorithms, study participants, and our laboratory staff for their contributions to this study.

## Author Contribution Statement

**Bechien U. Wu:** Conceptualization, Funding Acquisition, Investigation, Methodology, Project Administration, Resources, Supervision, Validation, Writing – Original Draft, Writing – Review & Editing.

**Tiffany Q. Luong:** Data Curation, Project Administration, Supervision, Visualization, Writing – Review & Editing

**Eva Lustigova:** Project Administration, Resources, Supervision, Writing – Review & Editing

**Christie Jeon:** Funding Acquisition, Investigation, Methodology, Writing – Review & Editing

**Eric J. Puttock:** Data Curation, Software, Writing – Review & Editing

**Rebecca H. Moon:** Data Curation, Validation, Writing – Review & Editing

**Mercedes A. Munis:** Data Curation, Project Administration, Supervision, Writing – Review & Editing

**Wansu Chen:** Conceptualization, Funding Acquisition, Investigation, Methodology, Project Administration, Resources, Supervision, Writing – Review & Editing

