## Supplemental File for "Implementation of risk prediction models using electronic health data for early detection of pancreatic cancer: a prospective pilot feasibility study"

**Supplementary File**

**Supplemental Figure 1.** Number of potentially eligible patients for each data pull.

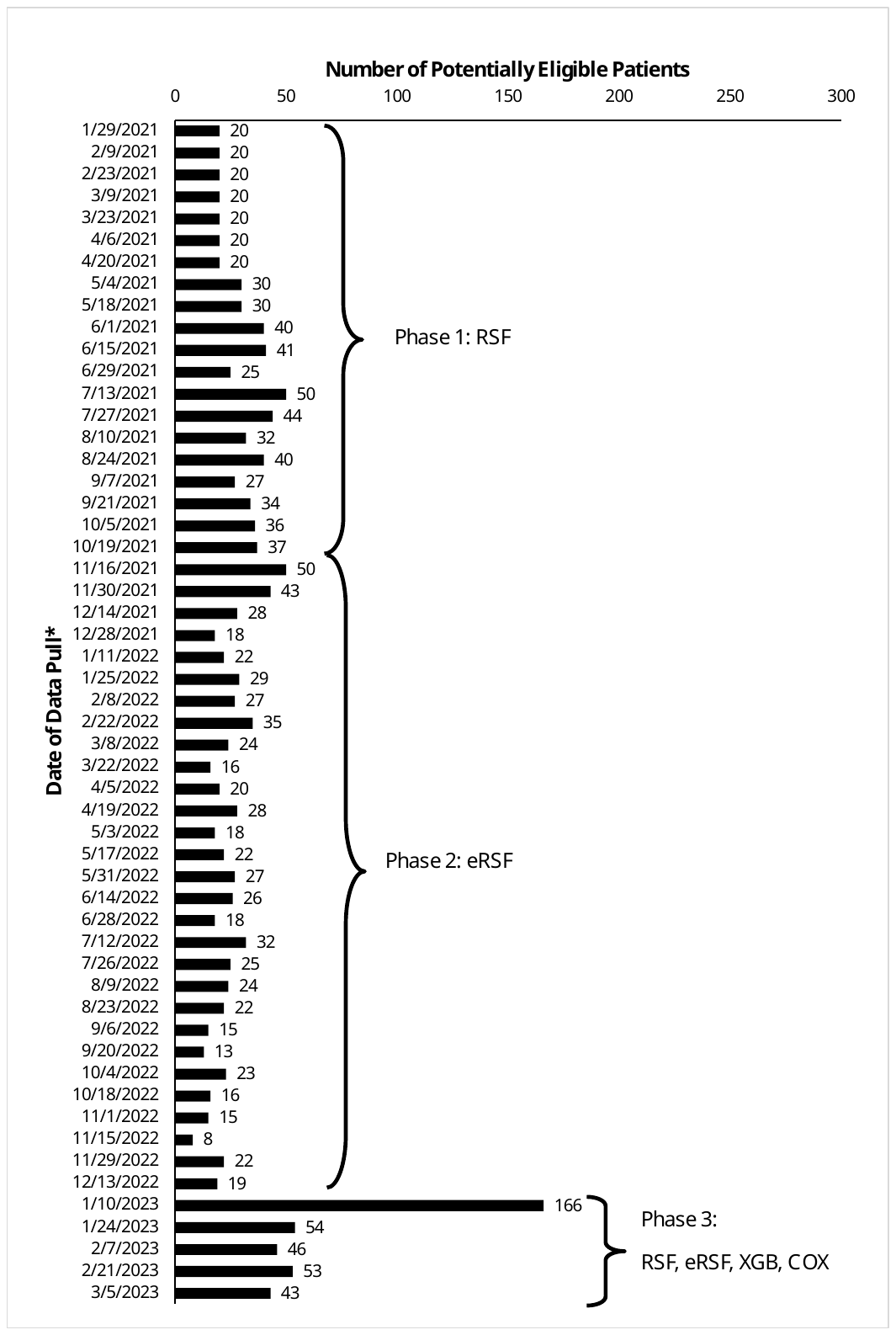

COX, Cox proportional hazards regression; eRSF, early detection random survival forest; RSF, random survival forest; XGB, extreme gradient boosting.

*A total of 54 data pulls were performed during the study. The Random Survival Forest (RSF) model was applied between January 29, 2021, and October 19, 2021. The early detection RSF model was applied between November 16, 2021, and December 13, 2022. All 4 models (RSF, eRSF, COX, and XGB) were applied between January 10, 2023, and March 05, 2023).

**Supplemental Figure 2.** Primary care physicians’ reasons for opting patient out from study participation.

|  | # Patients |
| --- | --- |
| Patient is not a good candidate | 3 |
| Patient is in hospice or palliative care | 2 |
| Patient has mental health issues | 2 |
| Patient is ill | 1 |
| Patient is bedbound | 1 |
| Patient is anxious | 1 |
| Patient is hard of hearing | 1 |
| Patient is planning to move outside study location | 1 |
| No reason provided | 8 |

**Supplemental Table 1.** Model parameters and performance (from Chen et al)

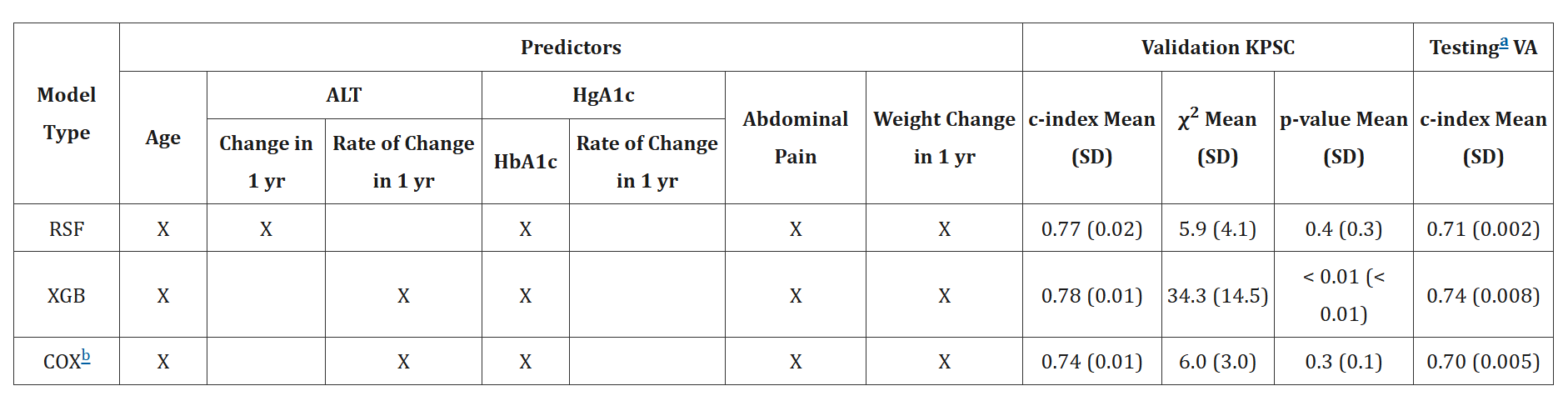

Model type: RSF: Random Survival Forest; XGB: eXtreme gradient boosting; COX: COX proportional hazards regression model.

Other abbreviations: ALT, alanine transaminase; C-index, concordance index; HgA1c, hemoglobin A1c; KPSC, Kaiser Permanente Southern California; NA, not applicable; SD, standard deviation; VA, Veterans Affairs

^a^Models were recalibrated using the exact features selected by RSF, XGB, and COX, respectively.

^b^The formula of the resulting prediction model is:

P(t) = P0(t) exp[βX], where βX = 0.073 × age + 1.012 × Abdominal Pain[yes] + 0.331 × Abdominal Pain[unknown] + 0.089 × HGBA1C – 0.055 × HGBA1C^2^ + 0.074 × [ALT Rate] – 0.001 × [ALT Rate]^2^ + 0.814 × [Weight Change] + 0.090 × [Weight Change]^2^

P(18 month) = 0.000052 × age + 0.000726 × Abdominal Pain[yes] + 0.000237 × Abdominal Pain[unknown] + 0.000064 × HGBA1C – 0.000039 × HGBA1C^2^ + 0.000053 × [ALT Change Rate] – 0.000001 × [ALT Change Rate]^2^ + 0.000584 × [Weight Change] + 0.000065 × [Weight Change]^2^

Reference: Chen W, Zhou B, Jeon CY, Xie F, Lin YC, Butler RK, Zhou Y, Luong TQ, Lustigova E, Pisegna JR, Wu BU. Machine learning versus regression for prediction of sporadic pancreatic cancer. Pancreatology. 2023 Jun;23(4):396-402. doi: 10.1016/j.pan.2023.04.009. Epub 2023 Apr 27. PMID: 37130760; PMCID: PMC10406388.

**Supplemental Table 2.** Classification of extra pancreatic findings.

| **Category** | **Description** |
| --- | --- |
| E1 | normal exam or normal variant |
| E2 | clinically unimportant finding, e.g. simple liver cyst, vertebral hemangioma: no work up required |
| E3 | likely unimportant or incompletely characterized finding, e.g. minimally complex renal cyst: referral depends on local center and standards of care |
| E4 | potentially important finding, e.g. solid renal mass, abdominal aortic aneurysm: communicate to referring physician |

**Supplemental Table 3.** Reasons for ineligibility (n=365).

| **Reason for Ineligibility** | **# Patients*** |
| --- | --- |
| Had cognitive impairment | 108 |
| Had active malignancy (other than pancreatic cancer), metastatic cancer, or was being treated with chemotherapy | 80 |
| Had metal or device (e.g., pacemaker) inside the body that precludes a patient from undergoing an MRI | 76 |
| Had cirrhosis | 60 |
| Had heart failure | 48 |
| Was in a skilled nursing facility or hospice care |  |
| Was hospitalized | 19 |
| Had eGFR <30 mL/min/1.73 m2 or end-stage renal disease | 18 |
| Was previously diagnosed with pancreatic cancer | 15 |
| Was deceased | 15 |
| Was unable to provide informed consent in either English or Spanish | 1 |
| Had health condition or physical limitation that prevented patient from participating in study activities | 1 |

eGFR, estimated glomerular filtration rate; MRI, magnetic resonance imaging.

*Numbers are not mutually exclusive. A patient may be ineligible according to multiple criteria.
